# How did exposure to Critical Access Hospitals change Medicare’s effect on cancer survival?

**DOI:** 10.1101/2025.03.10.25323691

**Authors:** Jason Semprini

**Affiliations:** Des Moines University College of Health Sciences, Department of Public Health

## Abstract

Upon turning 65, U.S. cancer patients experience the “Medicare Effect’ of better outcomes relative to patients nearly 65. Context likely mediates this effect, but so might differential access to care. With population-based cancer registry data, we construct a Difference-in-Discontinuities framework test if this “Medicare Effect” on cancer detection and survival varies in counties with and without a Critical Access Hospital (CAH), before and after CAH conversion. Consistent with prior literature, gaining Medicare was associated with reduced probability of distant stage diagnosis and improved two-year survival. Exposure to CAHs did not change Medicare’s effect on detection, but did increase Medicare’s effect on all-cause survival for males with prostate cancer (Est. = +0.019, se = 0.005), and females with lung (+0.046, se = 0.021) and colorectal cancer (Est. = +0.076, se = 0.026). Policies improving cancer survival remain warranted for both men and women with, and without Medicare.

## Introduction

Deemed the ‘Medicare Effect’, upon gaining Medicare coverage at the arbitrary age of 65 in the United States, health outcomes improve compared to adults nearly age 65[1], [2]. Under the seemingly justifiable assumption, that the only meaningful difference between adults just above and just below the age eligibility cutoff is enrollment in Medicare, access to Medicare causally impacts meaningful healthcare and health measures. Prior research, which almost uniformly has used a regression discontinuity design (RDD), has shown that Medicare, by reducing financial barriers to care[3], [4], [5], increases the utilization of preventative healthcare services[6], [7]. Additionally, the ‘Medicare Effect’ appears to increase utilization of post-acute, rehabilitation, and specialized services, including both high and low-value treatments[8], [9], [10], [11], [12], [13]. Effects are especially pronounced for adults requiring treatment for chronic health conditions[14], [15]. Greater access to care and lower financial strain may explain Medicare’s association with improved health status[16], [17], [18] and lower mortality[19].

This ‘Medicare Effect’, however, may not apply across all contexts, diseases, or populations. The discontinuities in healthcare or health outcomes are driven by disparate access to affordable health insurance between adults enrolled in Medicare and adults not yet eligible for Medicare[20], [21]. For example, as a result of increased spending and expanded health insurance coverage during the COVID-19 pandemic, gaining Medicare was not associated with excess mortality[22]. More generally, while recent research illuminated the impact of Medicare enrollment on early detection and mortality among cancer patients, this ‘Medicare Effect’ was only found in females[23]. Moreover, Myerson’s study focused solely on cancers with standardized screening protocols for this age group (female breast, colorectal, lung) and subsequent work illuminated that the ‘Medicare Effect’ may not manifest across all cancer sites or populations[23], [24]. One way Medicare could improve survival is through increased detection. In fact, Medicare has been found to increase receipt of mammograms[7], [23], colonoscopies[16], [23], and lung cancer screenings[25], which likely explains the associated increase in early-stage diagnoses[24], [25], [26]. Yet, the only study to examine Medicare’s causal effect on cancer mortality found that the change was driven by improved outcomes in females with lung cancer; suggesting that earlier detection, alone, may not fully explain Medicare’s positive impact on cancer mortality. However, while research has investigated how the ‘Medicare Effect’ may change in different contexts or policy environments, no research to our knowledge has assessed how Medicare’s effect on cancer survival changes amidst variation in access to healthcare services.

One of the population groups with no observed ‘Medicare Effect’ were rural Americans[24]. Despite consistent uptake in Medicare enrollment, gaining Medicare was not associated with earlier detection of any type of cancer prior to the health reforms of the 2010’s[24]. Consistent rural-urban disparities in screening and staging have been revealed for the three most common cancers with standardized screening protocols[27], [28], [29], [30]. Yet, these disparities cannot solely be due to insurance coverage, as the findings held even among rural-urban adults with similar private or public insurance[31], [32], [33]. Without dismissing the mechanisms linking Medicare coverage, early detection, and reduced cancer mortality; there is little evidence supporting the argument that the ‘Medicare Effect’ improves rural cancer outcomes through better detection. Rather, we observe a troubling trend: as the rural-urban gap in cancer detection narrows[34], [35], the rural-urban gap in cancer mortality widens[36].

Viewing rural-urban cancer disparities through a multilevel lens reveals the importance of local context and health system resources[37], [38], [39]. Regardless of rural status, most adults with cancer receive treatment at community hospitals, motivating efforts to increase, not just availability of detection services, but the quality of cancer treatment[40]. Over the past few decades, one of the most impactful events influencing access to and quality of care delivered by community cancer center hospitals was the proliferation of Critical Access Hospitals (CAHs)[41]. CAHs are critical assets for communities, most of which provide cancer screening and treatment services[42], [43]. Perhaps the emergence of CAHs, which may have impacted both the availability and quality of rural cancer services, influenced the ‘Medicare Effect’ on cancer detection and survivorship. Understanding how the protective effect of Medicare varies over time in environments with changing health system resources can illuminate how to improve access to quality of care for cancer patients both with, as well as without, Medicare coverage, while also displaying the possible consequences of failing to mitigate the persistent threat of closure or service elimination decisions facing community hospitals each year.

## Background on Critical Access Hospitals

While much of the research has emphasized the importance for patient care and health outcomes, Medicare has also proven to be critical for hospitals[44]. Policymakers have also relied heavily on Medicare as a tool for adapting and reforming the American health system[45]. One of such reforms was the Balanced Budget Act of 1997[46], which included provisions adapting Medicare hospital payment and delivery policies in response to the growing threat of hospital closures. Between 1990-2000, 460 hospitals closed[47]. Among the most important provisions were the Medicare Rural Hospital Flexibility (Flex) Program and the creation of a new hospital designation: the Critical Access Hospital (CAH). In brief, the reforms included adapting reimbursement schemes from prospective to cost-based and relaxed certain staffing requirements for specific hospitals in rural communities. Hospitals could receive these benefits by converting to a CAH if they met certain requirements (<25 acute inpatient beds, located 35 miles from another hospital, <96 hours average length of stay, provide 24/7 emergency services)[48]. Additionally, the Flex Program facilitated grant funds and technical capacity training for CAHs[48]. In 2023, there were 1,360 CAHs, which account for 22% of all U.S. hospitals[49].

While CAHs were designed to mitigate the risk of closure and maintain essential services for rural communities, questions remained about the scope and quality of services delivered by these facilities. While differences have attenuated over time, evidence suggested that observational care quality was lower in CAHs compared to other prospective payment hospitals[50]. Yet, there have been no reported differences between CAHs and non-CAHs of similar size in terms of quality or patient safety[51]. Specific to cancer services, over 75% of CAHs conduct blood testing, mammography, and/or colonoscopies[43]. However, there was considerable variation across states in the availability of cancer imaging and diagnostic services[52], [53], [54], [55].

While CAHs are less likely than larger hospitals to provide comprehensive cancer care, chemotherapy, and radiation therapy, CAHs are more likely to provide comprehensive cancer care and chemotherapy than similarly sized urban or rural prospective payment hospitals[56]. Over one third of all CAHs provide cancer surgery and chemotherapy, while only 5% of CAHs provide no cancer treatment. Despite these differences in availability of cancer screening and treatment services at CAHs, there has been no observed association between patient mortality and treatment at a CAH, at least for short-term outcomes[57], reemphasizing the potential role that CAHs may be serving in rural cancer control systems. Still, the limited evidence on quality of care hinders our understanding of how CAHs improve overall cancer survival. Moreover, given the intrinsic relationship between CAH and Medicare, it is unknown whether the designation of these facilities mitigated or exacerbated the observed disparities in cancer mortality between patients newly eligible and not yet eligible for Medicare.

## Methods

This study implements an emerging econometric approach that simultaneously estimates and compares four RDDs via a Differences-in-Discontinuities design[58], [59], [60].

### Conceptual Framework and Potential Outcomes

Conceptually, we are modelling discontinuities at age 65 for cancer patients in four mutually exclusive groups: counties without a CAH in the pre-period; counties without a CAH in the post-period, counties with a CAH in the pre-period; counties with a CAH in the post-period. Consider the instance where the ‘Medicare Effect’ remains unchanged after exposure to CAHs. This absence of a change could be driven by one of three events. First, there could very well be no ‘Medicare Effect’ on cancer survival for any county or time-period. Or, the absence of an observed change in the ‘Medicare Effect’ could be due to consistent changes in cancer outcomes for Medicare enrollees in both counties with and without a CAH. Or, even if there is a differential effect on cancer survival after exposure to CAHs, there could still be no observed change in the ‘Medicare Effect’ if outcomes improve similarly between Medicare and non-Medicare populations.

Alternatively, consider the result of an increasing ‘Medicare Effect’ after exposure to CAHs. This result, too, could be attributed to several potential outcomes. Imagine first, that exposure to CAHs, by increasing or maintaining access to care, increases Medicare’s effect on cancer outcomes. Conversely, this same result could be due to declining outcomes as a result of less access to comprehensive care for non-Medicare enrollees. Or, finally, even if there is no observed change in the ‘Medicare Effect’ among cancer patients after exposure to CAHs, the ‘Medicare Effect’ could increase depending on the national trends in cancer outcomes for adults enrolled and not-yet-enrolled in Medicare. If outcomes improve for non-Medicare enrollees, nationally, but not in counties with a CAH, that will yield a result of an increasing ‘Medicare Effect’ after exposure to CAHs. The counter to these hypothetical potential outcomes holds for the third result: that exposure to CAH’s decreases the ‘Medicare Effect’. The ‘Medicare Effect’ could decrease due to worsening quality for Medicare enrollees, improving access for non-enrollees, or a stagnate ‘Medicare Effect’ in CAH counties amidst a rising ‘Medicare Effect’ nationally.

### Data and Sample

Data for this study was obtained from the Surveillance, Epidemiology, and End Results (SEER) program’s SEER-13 datafile (1992-2020)[61]. SEER collects a census of cancer cases diagnosed within participating registries. This population-based data represents the U.S. cancer burden and accounts for nearly half of all cancers diagnosed in the U.S. each year. The SEER-13 datafile includes thirteen participating registries. For this study, the Alaska Native Tumor Registry and Hawaii’s registry were excluded, leaving ten registries in eight states: Connecticut, Georgia (Atlanta Registry, Rural Georgia Registry), California (San-Francisco-Oakland Registry, San Jose-Monterey Registry, Los Angeles Registry), Iowa, Michigan (Detroit Registry), New Mexico, Washington (Seattle-Puget Sound Registry), and Utah. For each cancer case, SEER includes data on tumor characteristics, individual sociodemographic information, and survival. As of November 2022, SEER-13 also included state and county geographic identifiers which were merged at the county-level with historic CAH data from the Flex Monitoring Team[62].

In addition to excluding data from Alaska and Hawaii, this study also excludes data from years 1999-2005. Considered a ‘wash out’ period, it was during this time period that states began participating in the Flex Program and converting hospitals to CAHs. Additionally, the study included only individuals between ages 45-84 who have been diagnosed with a first-malignant primary tumor (diagnostic confirmation). This inclusion criteria excludes cancer patients farther from the age 65 cutoff, patients with multiple tumors, and cancers discovered during an autopsy or during end-of-life care.

### Measures and Subgroup Analyses

The two primary outcomes measure staging at diagnosis and patient survival. Using SEER’s Historic Summary Stage A variable[63], the first outcome was modelled as a binary variable indicating if the patient was diagnosed at a distant stage. Unstaged cancers were excluded. The second outcome uses SEER derived survival data to construct a binary variable indicating if the patient survived two-or-more years from diagnosis (all-cause survival). The primary exposure variable was being age 65 or older at diagnosis. SEER’s sociodemographic data on sex, race/ethnicity, and marital status were used to construct a set of binary control variables (Table 1). In addition to analyzing the full sample of cancer cases meeting the inclusion criteria, this study also conducts subgroup analyses for four high-incidence cancer sites: lung, prostate, female breast, colorectal[64]. Cancer sites were identified by the SEER derived ICD-03 recode variable. For all sites combined, lung, and colorectal, this study also conducted subgroup analyses by sex.

**Table 1:** Sample Statistics. reports the summary statistics of the analytic sample. Each row measures the proportion of the sample of that specific variable for each time (pre/post) and county (CAH, non-CAH).

|  |  | 1993-1998 |  | 2006-2010 |  |
| --- | --- | --- | --- | --- | --- |
|  |  | None | CAH | None | CAH |
| <u>**Outcomes**</u> | Distant | 24.6% | 25.8% | 22.7% | 24.1% |
|  | Two-Year Survival | 66.2% | 65.2% | 72.8% | 71.0% |
| <u>**Age**</u> | Age (continuous) | (66.6) | (67.0) | (65.0) | (65.3) |
|  | Medicare | 60.5% | 62.0% | 50.7% | 52.6% |
| <u>**Sociodemographics**</u> | Female | 45.6% | 45.6% | 45.4% | 46.0% |
|  | Male | 54.4% | 54.4% | 54.6% | 54.0% |
|  | Hispanic | 5.1% | 10.0% | 7.5% | 14.7% |
|  | non-Hispanic AIAN | 0.4% | 0.3% | 0.6% | 0.5% |
|  | non-Hispanic API | 3.8% | 6.4% | 5.9% | 9.4% |
|  | non-Hispanic Black | 10.9% | 8.5% | 12.2% | 8.3% |
|  | non-Hispanic unknown race | 0.4% | 0.6% | 0.9% | 1.1% |
|  | non-Hispanic White | 79.4% | 74.3% | 73.0% | 66.0% |
|  | Divorced | 8.1% | 8.8% | 10.1% | 9.6% |
|  | Married | 59.8% | 60.5% | 56.5% | 56.4% |
|  | Separated | 0.6% | 0.7% | 0.9% | 0.9% |
|  | Never Married | 9.5% | 10.0% | 13.1% | 13.9% |
|  | Unknown | 5.5% | 3.7% | 8.4% | 8.0% |
|  | Widowed | 16.5% | 16.3% | 11.1% | 11.1% |
| <u>**Cancer Sites**</u> | Lung | 14.7% | 14.2% | 12.1% | 11.5% |
|  | Prostate | 19.7% | 19.3% | 20.8% | 18.9% |
|  | Female Breast | 14.9% | 14.7% | 14.5% | 14.8% |
|  | Colorectal | 10.0% | 10.2% | 8.2% | 9.0% |
| <u>**States**</u> | California | 27.1% | 59.5% | 26.4% | 57.9% |
|  | Connecticut | 18.9% | 0.0% | 17.4% | 0.0% |
|  | Georgia | 9.6% | 0.4% | 11.4% | 0.7% |
|  | Iowa | 6.2% | 16.1% | 6.0% | 14.6% |
|  | Michigan (Detroit) | 22.1% | 0.0% | 19.0% | 0.0% |
|  | New Mexico | 6.3% | 1.0% | 7.5% | 1.0% |
|  | Utah | 5.7% | 0.5% | 7.2% | 0.6% |
|  | Washington (Seattle-Puget Sound) | 4.2% | 22.5% | 5.2% | 25.2% |
| <u>**Rurality**</u> | Metro | 92.1% | 82.1% | 93.4% | 85.1% |
|  | Urban | 7.3% | 15.7% | 6.2% | 13.1% |
|  | Rural | 0.6% | 2.2% | 0.4% | 1.8% |
| **N** |  | 400,617 | 232,961 | 391,448 | 220,926 |

### Design

Two commonly used quasi-experimental designs, the Difference-in-Differences (DD) and Regression Discontinuity Design (RDD) serve as the foundation for this study’s Difference-in-Discontinuities Design[58], [59], [60]. Readers may be familiar with the first two common designs, but less so with the Difference-in-Discontinuities design. However, the intuition is quite clear and straightforward. To see this, first consider a canonical DD with a single post period indicator (POST_t_), which equals 1 if the year = t is between 2006-2010 and zero otherwise, and a single treatment indicator (CAH_c_), which equals 1 if the county = c has a CAH and zero otherwise.

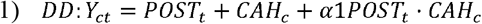

Equation 1 calculates the relative difference in cancer outcome trends between counties with, and without, a CAH, between the two time periods. For valid identification, the α1 parameter, the association between exposure to a CAH and cancer outcomes, must satisfy the independence of potential outcomes assumption, often referred to as the parallel trends assumption. This identification strategy assumes the trends in counties with a CAH (c=1) would have remained parallel to counties without a CAH (c=0) in the absence of CAH exposure (Equation 2). This assumption may prove difficult to justify, given that the introduction of a CAH was not random, but in fact quite plausibly related to the health status and potential mortality outcomes of its nearby residents. Failing to satisfy the parallel trends assumption (Equation 2) will yield biased DD estimates.

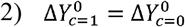

While exposure to a CAH hospital may be endogenous with cancer outcomes, the second part of our design does not suffer from such a flagrant threat to validity. The vast literature on the ‘Medicare Effect’ posits that the potential outcomes of an individual newly eligible for Medicare are independent of potential outcomes of an individual nearly, but not yet, eligible for Medicare; and the only differences observed between these two individuals will be related to Medicare enrollment (Equation 3).

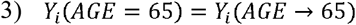

To estimate the ‘Medicare Effect’ (α2 in Equation 4) using an RDD, the model must include an indicator for Medicare status (MEDCARE_i_), which takes a value of 1 if the individual is age 65 or above, and a set of slope parameters which measure the association between age and cancer outcomes for non-Medicare enrollees and Medicare enrollees (Equation 4).

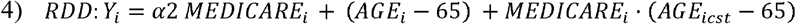

Conceptually, a Difference-in-Discontinuities is simply estimating Equation 4 among four groups: a) POST=0, CAH = 0; b) POST = 1, CAH = 0; c) POST = 0, CAH = 1; d) POST = 1, CAH = 1. Equation 5 shows how Equation 1 and Equation 3 combine into a fully saturated Difference-in-Discontinuities model.

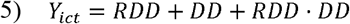

For valid inference, Equation 5 no longer requires parallel trends or independence of potential outcomes between cancer patients exposed to a CAH and cancer patients not exposed to a CAH. Rather, Equation 5 only requires that any bias caused by differential trends between CAH and non-CAH counties is similar for adults at or near the Medicare eligibility cutoff in each group G = (a, b, c, d). As shown in Equation 3, we are assuming that potential outcomes are independent of Medicare eligibility near age 65. Under this, again seemingly valid and well supported assumption, we extend the implications of Equation 3 to independence of potential bias:

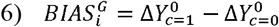

Equation 6 measures the bias resulting from the non-common trends of exposure to a CAH for individuals = i, for each of the four RDD groups = G (a, b, c, d). Substituting this bias measure into Equation 3, we now assume that the potential bias, for each RDD group = G, is similar between adults just age 65 and adults nearly 65 (Equation 7).

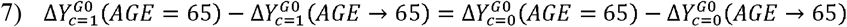

Under the new identification assumption in Equation 7, we model our two cancer outcomes as a fully saturated Difference-in-Discontinuities model. The primary goal of Equation 8 is to identify β1, the change in the ‘Medicare Effect’ on cancer outcomes after exposure to a CAH.

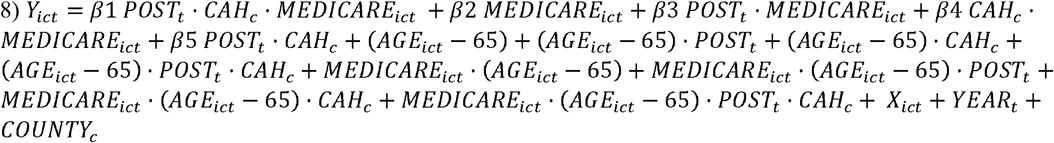

For the distant stage analyses, Y_ict_ measures the probability of being diagnosed at a distant stage. For survival analyses, Y_ict_ measures the probability of surviving two years. In both cases, the outcome is a probability measure for each individual = i, diagnosed in year = t and county = c. The β2 parameter adjusts for the overall ‘Medicare. The β3 parameter adjusts for the how the ‘Medicare Effect’ changes between 1993-1999 to 2006-2010. The β4 parameter adjusts for the baseline ‘Medicare Effect’ in counties with a CAH. The β5 parameter adjusts for the association between exposure to CAHs and cancer outcomes. Equation 8 also includes a fully saturated set of slope parameters adjusting the for association between age and outcomes. Additionally, Equation 8 adjusts for sociodemographic differences between individuals (X_ict_’), temporary trends with year fixed-effects (YEAR_t_), and baseline county-level differences with county-level fixed-effects (COUNTY_c_).

### Analyses

All analyses were estimated via a linear probability regression model in STATA v. 18[65]. For inference, standard errors robust to heteroskedasticity were clustered at the county-level[66]. To assess if the results were sensitive to model specification, the above analyses were repeated by 1) including a state-year interaction fixed-effect to account for time-variant differences in cancer outcomes at the state-level and 2) adding a saturated set of quadratic age slope terms[67]. To test the validity of our identification strategy and design, we re-estimate the above models with a placebo treatment variable (cutoff = age 55, cutoff = age 75).

## Results

The analytic sample include 1,245,952 adults with cancer. Table 1 reports the sample average for both outcomes, age, Medicare eligibility, sociodemographic control variables, cancer sites, and state registries. Supplemental Exhibit 1 reports the cancer and sex-specific analytic counts, and the sample average for adults age 65 for both outcome variables. Overall, 23.5% of the sample were diagnosed at a distant stage and 72.2% of the sample survived two-years (Supplemental Exhibit 1). See the online appendix for the full set of results for each model, including the coefficients and standard errors for all model parameters. When combining all cancers and counties, there is a clear discontinuity at age 65 with an associated decline in the probability of a distant stage diagnosis and an associated increase in the probability of two-year survival (Supplemental Exhibit 2).

## Difference-in-Discontinuities Estimates

### Distant Stage

There is little convincing evidence that exposure to CAHs changed the ‘Medicare Effect’ on cancer detection. For all sites combined, the interaction between Medicare eligibility and exposure to a CAH was not associated with the probability of a distant stage diagnosis (Est. = +0.004, se = 0.006; Table 2; Supplemental Exhibit 3). Consistent with the overall estimate, the interaction between Medicare eligibility and exposure to a CAH on the probability of a distant stage diagnosis was near zero and statistically insignificant for lung and prostate cancers (Table 1, Supplemental Exhibit 4). We do, however, find that the effect of Medicare on the probability of a distant stage cancer increased after exposure to CAHs for all cancer sites combined in females (Est. = +0.016, se = 0.008, p < 0.05; Table 1). This estimate may be related to the statistically significant increase for female breast cancers, where the ‘Medicare Effect’ is associated with lower probability of distant stage diagnoses in counties with a CAH before conversion, but less so after conversion (Supplemental Exhibit 4). Finally, the interaction between Medicare eligibility and exposure to a CAH is associated with a decrease in the probability of a distant stage diagnosis for colorectal cancer cases (Est. = −0.035, se = 0.016, p < 0.05; Table 1). While there are no differences by sex, the estimate is no longer statistically significant when including the quadratic age slope (Supplemental Exhibit 5).

**Table 2:** Difference-in-Discontinuities Estimates. reports the effect estimates of the Difference-in-Discontinuities, estimating how exposure to county-level CAH conversion changed the Medicare Effect on each outcome. Standard errors reported in parentheses. * p < 0.05, ** p < 0.01, *** p < 0.001

|  | <u>Distant Stage</u> |  |  | <u>Two-Year Survival</u> |  |  |
| --- | --- | --- | --- | --- | --- | --- |
|  | **Both** | **Female** | **Male** | **Both** | **Female** | **Male** |
| **<u>All Sites</u>** |  |  |  |  |  |  |
| *Est.* | 0.004 | 0.016* | -0.007 | 0.002 | -0.005 | 0.008 |
| *(se)* | (0.006) | (0.008) | (0.007) | (0.005) | (0.007) | (0.008) |
| **<u>Lung</u>** |  |  |  |  |  |  |
| *Est.* | -0.003 | 0.008 | -0.010 | 0.019 | 0.046* | -0.003 |
| *(se)* | (0.021) | (0.029) | (0.022) | (0.015) | (0.021) | (0.020) |
| **<u>Colorectal</u>** |  |  |  |  |  |  |
| *Est.* | -0.035* | -0.036 | -0.034 | 0.050** | 0.076** | 0.031 |
| *(se)* | (0.016) | (0.022) | (0.021) | (0.016) | (0.026) | (0.020) |
| **<u>Female Breast</u>** |  |  |  |  |  |  |
| *Est.* | - | 0.014 | - | - | -0.017* | - |
| *(se)* | - | (0.008) | - | - | (0.008) | - |
| **<u>Male Prostate</u>** |  |  |  |  |  |  |
| *Est.* | - | - | 0.001 | - | - | 0.019*** |
| *(se)* | - | - | (0.007) | - | - | (0.005) |

### Two-Year Survival

For all cancer sites combined, the interaction between Medicare eligibility and CAH exposure was not associated with the probability of two-year survival (Est. = +0.002, se = 0.005; Table 2; Figure 1). There were no sex differences. The estimated interaction between Medicare eligibility and CAH exposure on two-year survival of lung cancer was also statistically insignificant (Est. = +0.019, se = 0.015; Table 1; Figure 2). However, when including the quadratic age slopes, the estimated interaction between Medicare eligibility and CAH exposure on two-year survival of lung cancer was larger in magnitude and statistically significant (Est. = +0.058, se = 0.023, p < 0.05; Supplemental Exhibit 6). Only for lung cancer survival was there any meaningful difference between the primary estimate and the estimate with quadratic age slopes (Supplemental Exhibit 6). Regarding sex differences, while not statistically different, there were meaningful differences between females and males for the estimated interaction between Medicare eligibility and exposure to CAHs on two-year survival in lung cancer (Females: Est. = +0.046, se = 0.021, p < 0.05; Males: = −0.003, se = 0.020; Table 2).

**Figure 1:**
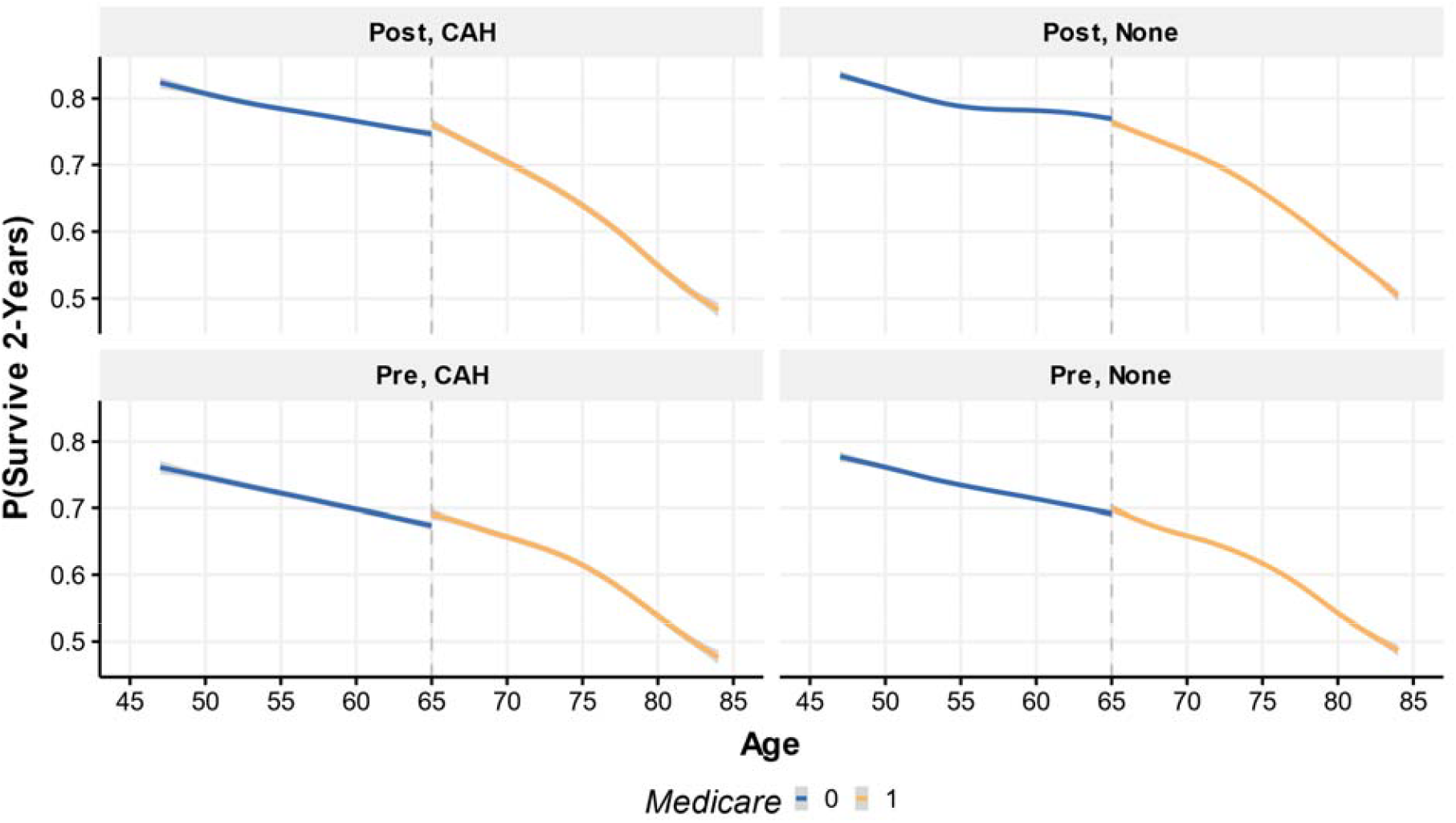
Differences-in-Discontinuities of Two-Year Survival (All Sites Combined) visualizes the discontinuity in the probability of two-year survival among adults at age 65 (all cancer sites combined). Lines represent the average probability at each age using locally estimated scatterplot smoothing method. Shaded gray area represents 95% Confidence Interval. Pre = 1993-1998, Post = 2006-2010. None = county did not have a CAH, CAH = county had at least one CAH.

**Figure 2:**
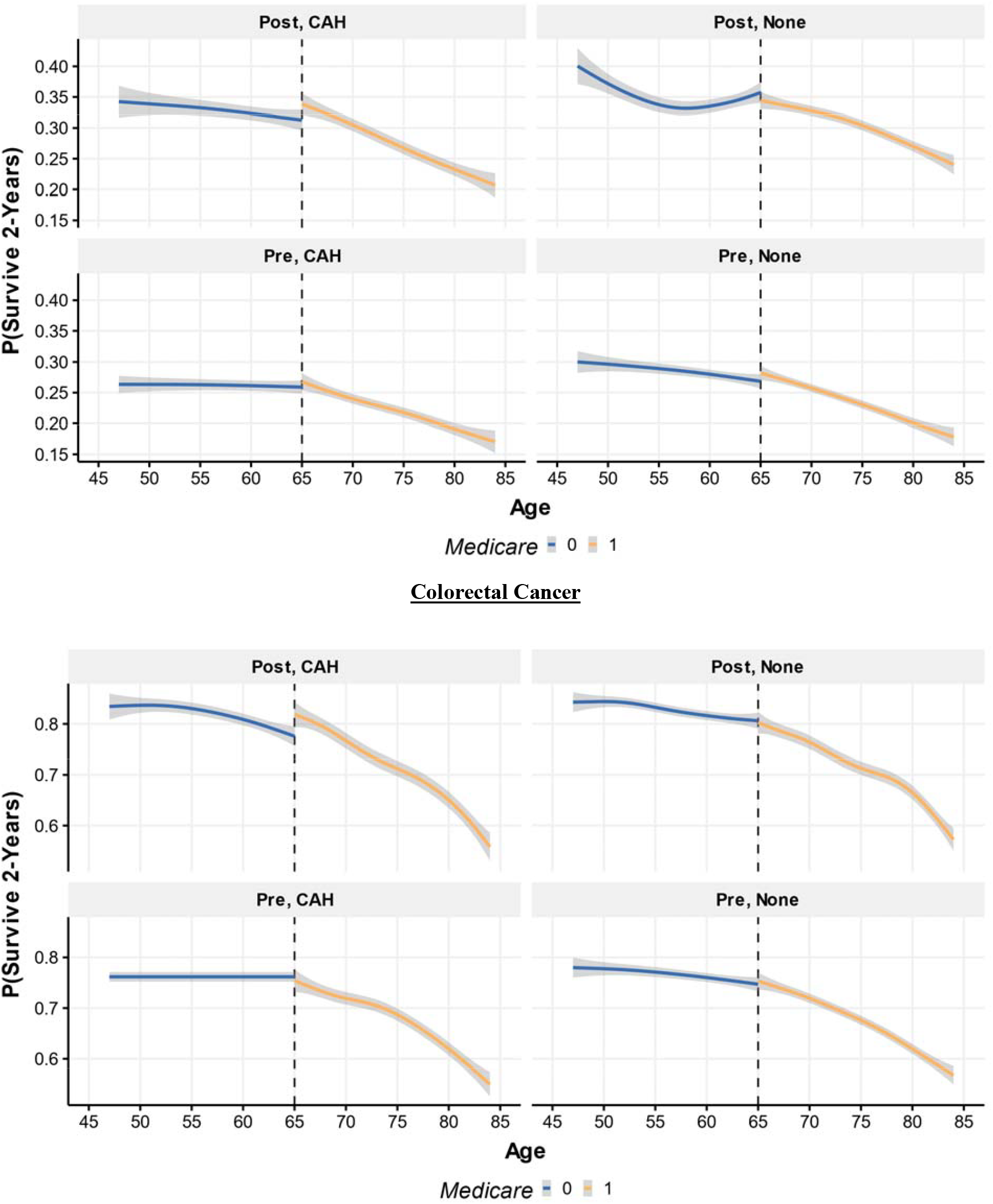
Differences-in-Discontinuities of Two-Year Survival (Lung & Colorectal) visualizes the discontinuity in the probability of two-year survival among adults with lung cancer and colorectal cancer at age 65. Lines represent the average probability at each age using locally estimated scatterplot smoothing method. Shaded gray area represents 95% Confidence Interval. Pre = 1993-1998, Post = 2006-2010. None = county did not have a CAH, CAH = county had at least one CAH.

Figure 3 visualizes the discontinuity at age 65 for two-year survival for female breast cancer and prostate cancer. For female breast cancer, there was a statistically significant negative association between the interaction of Medicare eligibility and CAH exposure with two-year survival (Est. = −0.017, se = 0.008, p < 0.05). The statistical significance of this estimate, however, attenuates with the quadratic age slope specification. For prostate cancer, the interaction between Medicare eligibility CAH exposure with two-year survival was statistically significant and positive (Est. = +0.019, se = 0.005, p < 0.001; Table 2). This estimate is consistent across the primary, quadratic age slope, and state-year fixed effect specifications, and corresponds to a 2% relative increase in the probability of two-year survival of prostate cancer.

**Figure 3:**
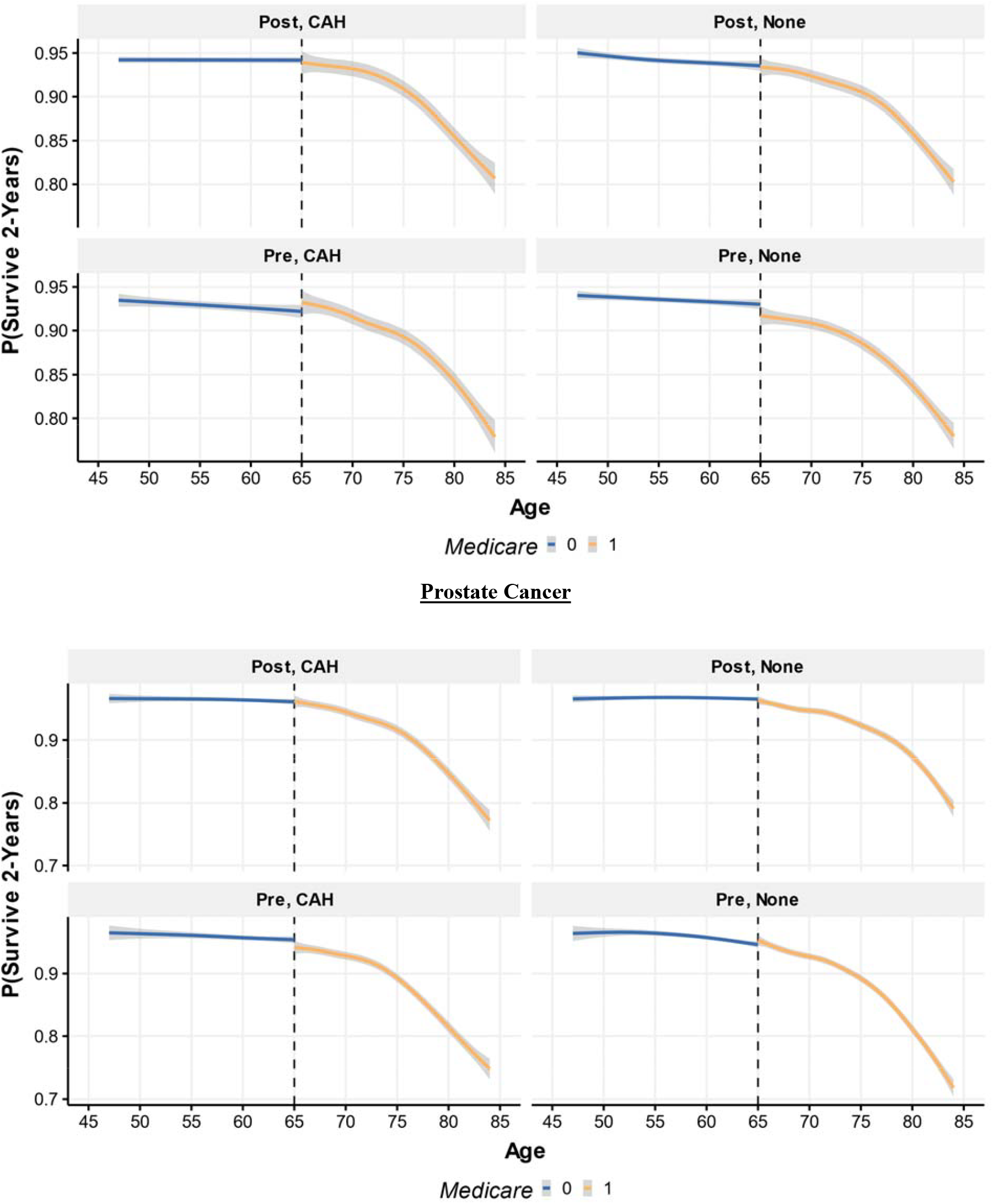
Differences-in-Discontinuities of Two-Year Survival (Female Breast & Prostate) visualizes the discontinuity in the probability of a two-year survival among adults with female breast and prostate cancer at age 65. Lines represent the average probability at each age using locally estimated scatterplot smoothing method. Shaded gray area represents 95% Confidence Interval. Pre = 1993-1998, Post = 2006-2010. None = county did not have a CAH, CAH = county had at least one CAH.

Finally, the interaction between Medicare eligibility and CAH exposure is associated with statistically significant increase in the probability of two-year survival of colorectal cancer (Est. = 0.050, se = 0.016, p < 0.01; Table 1; Figure 2). This estimate is consistent across model specifications and corresponds to a 6.5% relative increase. While not statistically different, again there appear to be meaningful differences by sex, with the estimate for females (Est. = 0.076, se = 0.026, p < 0.001) more than double the estimate for males (Est. = 0.031, se = 0.020). The female estimate corresponds to a 10% relative increase in the probability of surviving two years after diagnosis.

## Discussion

This study examined how the phenomenon known as the ‘Medicare Effect’ varied by exposure to CAHs. Consistent with prior work, there was a clear discontinuity in survival at age 65 overall, but only for specific cancers and with larger effects in females[23], [24]. CAH exposure was not consistently found to significantly change in Medicare’s effect on early detection. There was also little consistent evidence that CAH exposure changed Medicare’s effect on two-year survival. Aside from the small relative increase in survival for males with prostate cancer, the only consistent, significant, and clinically meaningful differences from Medicare’s effect after exposure to CAHs on two-year survival was for females with lung or colorectal cancer.

For decades, mortality rates in these two common cancers have declined[68], [69], [70]. The improved survival, observed across much of the population, including both males and females, has been largely attributed to improved treatment[69]. Gaining Medicare eligibility at age 65 after exposure to CAHs may have led to improved survival among females with lung or colorectal cancer because of the availability of healthcare services. These effects can, at most, only partially be explained by earlier detection, which is consistent with the evidence showing improved detection amidst worse survival in rural cancer patients[34], [35], [36]. For colorectal cancer, the estimated change in survival exceeds the estimated change in distant stage diagnoses and for lung cancer, the change in distant stage diagnoses was near zero. Rather, the observed survival improvements upon gaining Medicare after exposure to CAH is more likely to be due to improved or maintained access to high-quality healthcare services.

Colorectal cancer is among the most common cancers treated at CAHs[43], so it should be no surprise that we observe a differential improvement in survival for Medicare enrollees after exposure to CAHs given the importance of hospital volume[71]. It is less likely, however, that the differential survival benefit for Medicare enrollees after exposure to CAHs was due to increased or maintained access to lung cancer treatment, as CAHs treat fewer lung cancer patients and have less capacity to deliver multidisciplinary care[43], [72]. However, because we measure all-cause survival, and not cancer specific survival, the improved survival benefit at the age of 65 after exposure to CAHs could possibly be explained by greater availability of healthcare services, generally. Future research should further examine these possible mechanisms to inform ongoing cancer control policies. Innovative models for delivering comprehensive cancer care to patients at CAHs could further improve cancer outcomes.

While beneficial to Medicare beneficiaries, from a societal perspective we should be applaud a diminishing a ‘Medicare Effect’. Disparities in cancer survival at the arbitrary age of 65 reflect broader disparities within our healthcare system. For many of the cancers analyzed in this study, the discontinuity in survival at age 65 attenuated and in some instances, disappeared between 1998 and 2010. Many factors could explain this, including the growing proliferation of Medicare Advantage or cancer-specific Medicaid eligibility insurance programs[73], [74]. Looking ahead, we predict even less of a ‘Medicare Effect’ on cancer survival, given the Affordable Care Act’s impact on increased access to and utilization of preventative and cancer treatment services[75], [76]. As the U.S. continues to pursue a more equitable healthcare system and universal insurance coverage, we should expect a continued diminishing importance of the ‘Medicare Effect’.

Still, Medicare, as insurance coverage and a policymaking tool, remains important for adults with cancer, just as Critical Access Hospitals remain critical features of rural cancer control systems. As rural hospitals continue to face financial pressure to close, eliminate service lines, or redesignate to alternative models of delivery[77], [78], policymakers must continue to monitor, evaluate, and implement policies to ensure adequate access to high-quality cancer care. This study presents evidence that exposure to CAHs may have been associated with improved survival for Medicare enrollees. This observed benefit, which was not experienced by males or adults with less common cancers, is driven by the discontinuities in access to insurance and lower propensity to access high-quality care. Through insurance reform and health-system innovation, our goal as policymakers and researchers should be to eliminate these discontinuities.

## Limitations

Since the data was accessed in Nov. 2022, the Detroit registry and county-level identifiers were removed from the public SEER datafiles. An additional data limitation stems from the lack of insurance information in the public SEER datafile. Moreover, the RDD continuity assumption could be improved with month of diagnosis and month of birth data. Also, the limited availability of treatment data in the public SEER file hinder any attempt to test the mechanisms of our results. Finally, as with all quasi-experimental designs analyzing retrospective, observational data, the study’s ability to infer causality remains limited by the validity of our stated assumptions. As the Difference-in-Discontinuities design becomes more widely utilized and assessed empirically, as a field we will be better equipped to diagnose and discuss the strengths and weaknesses of this emerging methodology.

## Conclusions

Upon turning 65, cancer patients experience a “Medicare Effect” leading to improved outcomes relative to patients nearly 65. Using population-based data, this study was among the first to assess how this ‘Medicare Effect’ varies with differential access to healthcare services as measured by exposure to a CAH. While there was little evidence to support that the effect of Medicare on earlier detection varied by CAH exposure, there was consistent evidence that Medicare’s positive impact on survival increased with exposure to CAHs for females with lung and colorectal cancers, and males with prostate cancer. This study reiterates the importance of, not just insurance, but access to healthcare services. As the ‘Medicare Effect’ continues to change over time, policies ensuring adequate access to high-quality cancer care remain warranted, for adults with, and without, Medicare coverage.

## Data Availability

Data is sharing subject to third-party restrictions (NCI).

## Notes

### Competing Interest Statement

The authors have declared no competing interest.

### Funding Statement

This study did not receive any funding.

### Summary of Updates

Updated reference list and style.

## References

[1] H. Levy and D. Meltzer, “The Impact of Health Insurance on Health,” Annual Review of Public Health, vol. 29, no. 1, pp. 399–409, 2008, doi: 10.1146/annurev.publhealth.28.021406.144042.

[2] D. Card, C. Dobkin, and N. Maestas, “The Impact of Nearly Universal Insurance Coverage on Health Care Utilization: Evidence from Medicare,” Am Econ Rev, vol. 98, no. 5, pp. 2242–2258, Dec. 2008, doi: 10.1257/aer.98.5.2242.

[3] E. T. Roberts, Y. Kwon, A. G. Hames, J. M. McWilliams, J. Z. Ayanian, and R. Tipirneni, “Racial and Ethnic Disparities in Health Care Use and Access Associated With Loss of Medicaid Supplemental Insurance Eligibility Above the Federal Poverty Level.,” JAMA Intern Med, vol. 183, no. 6, pp. 534–543, Jun. 2023, doi: 10.1001/jamainternmed.2023.0512.

[4] P. U. Neiman, V. H. Mouli, K. K. Taylor, J. Z. Ayanian, J. B. Dimick, and J. W. Scott, “The Impact of Medicare Coverage on Downstream Financial Outcomes for Adults Who Undergo Surgery.,” Ann Surg, vol. 275, no. 1, pp. 99–105, Jan. 2022, doi: 10.1097/SLA.0000000000005272.

[5] R. Aggarwal, R. W. Yeh, I. J. Dahabreh, S. E. Robertson, and R. K. Wadhera, “Medicare eligibility and healthcare access, affordability, and financial strain for low- and higher-income adults in the United States: A regression discontinuity analysis.,” PLoS Med, vol. 19, no. 10, p. e1004083, Oct. 2022, doi: 10.1371/journal.pmed.1004083.

[6] J. M. McWilliams, A. M. Zaslavsky, E. Meara, and J. Z. Ayanian, “Impact of Medicare coverage on basic clinical services for previously uninsured adults,” JAMA, vol. 290, no. 6, pp. 757–764, Aug. 2003, doi: 10.1001/jama.290.6.757.

[7] P. D. Jacobs and S. Abdus, “Changes in preventive service use by race and ethnicity after medicare eligibility in the United States.,” Prev Med, vol. 157, p. 106996, Apr. 2022, doi: 10.1016/j.ypmed.2022.106996.

[8] S. E. Regenbogen, A. H. Cain-Nielsen, J. D. Syrjamaki, L. M. Chen, and E. C. Norton, “Spending On Postacute Care After Hospitalization In Commercial Insurance And Medicare Around Age Sixty-Five.,” Health Aff (Millwood), vol. 38, no. 9, pp. 1505–1513, Sep. 2019, doi: 10.1377/hlthaff.2018.05445.

[9] C. K. Zogg et al., “The Association Between Medicare Eligibility and Gains in Access to Rehabilitative Care: A National Regression Discontinuity Assessment of Patients Ages 64 Versus 65 Years.,” Ann Surg, vol. 265, no. 4, pp. 734–742, Apr. 2017, doi: 10.1097/SLA.0000000000001754.

[10] E. Lin, G. M. Chertow, J. Bhattacharya, and D. Lakdawalla, “Early Delays in Insurance Coverage and Long-term Use of Home-based Peritoneal Dialysis.,” Med Care, vol. 58, no. 7, pp. 632–642, Jul. 2020, doi: 10.1097/MLR.0000000000001350.

[11] S. Park, R. K. Wadhera, and J. Jung, “Effects of Medicare eligibility and enrollment at age 65?years on the use of high-value and low-value care.,” Health Serv Res, vol. 58, no. 1, pp. 174–185, Feb. 2023, doi: 10.1111/1475-6773.14065.

[12] A. C. De Roo, J. Ha, S. E. Regenbogen, and G. J. Hoffman, “Impact of Medicare eligibility on informal caregiving for surgery and stroke.,” Health Serv Res, vol. 58, no. 1, pp. 128–139, Feb. 2023, doi: 10.1111/1475-6773.14019.

[13] J. Dugan, S. S. Virani, and V. Ho, “Medicare eligibility and physician utilization among adults with coronary heart disease and stroke.,” Med Care, vol. 50, no. 6, pp. 547–553, Jun. 2012, doi: 10.1097/MLR.0b013e318245a64d.

[14] J. H. Rhodes, “Changes in the Utilization of Mental Health Care Services and Mental Health at the Onset of Medicare.,” J Ment Health Policy Econ, vol. 21, no. 1, pp. 29–41, Mar. 2018.

[15] B. J. Lipton, “Association between Health Insurance and Health among Adults with Diabetes: Evidence from Medicare.,” J Am Geriatr Soc, vol. 68, no. 2, pp. 388–394, Feb. 2020, doi: 10.1111/jgs.16238.

[16] S. Park, A. N. Ortega, J. Chen, and A. V. Bustamante, “Effects of Medicare eligibility and enrollment at age 65 among immigrants and US-born residents.,” J Am Geriatr Soc, Apr. 2023, doi: 10.1111/jgs.18380.

[17] D. Polsky et al., “The health effects of Medicare for the near-elderly uninsured,” Health Serv Res, vol. 44, no. 3, pp. 926–945, Jun. 2009, doi: 10.1111/j.1475-6773.2009.00964.x.

[18] J. Wallace, K. Jiang, P. Goldsmith-Pinkham, and Z. Song, “Changes in Racial and Ethnic Disparities in Access to Care and Health Among US Adults at Age 65 Years.,” JAMA Intern Med, vol. 181, no. 9, pp. 1207– 1215, Sep. 2021, doi: 10.1001/jamainternmed.2021.3922.

[19] D. Card, C. Dobkin, and N. Maestas, “Does Medicare Save Lives?*,” The Quarterly Journal of Economics, vol. 124, no. 2, pp. 597–636, May 2009, doi: 10.1162/qjec.2009.124.2.597.

[20] M. R. Weaver et al., “Variation in Health Care Access and Quality Among US States and High-Income Countries With Universal Health Insurance Coverage,” JAMA Netw Open, vol. 4, no. 6, p. e2114730, Jun. 2021, doi: 10.1001/jamanetworkopen.2021.14730.

[21] S. A. Lavarreda, E. R. Brown, and C. D. Bolduc, “Underinsurance in the United States: an interaction of costs to consumers, benefit design, and access to care,” Annu Rev Public Health, vol. 32, pp. 471–482, 2011, doi: 10.1146/annurev.publhealth.012809.103655.

[22] J. Wallace, A. Lollo, and C. D. Ndumele, “Evaluation of the Association Between Medicare Eligibility and Excess Deaths During the COVID-19 Pandemic in the US.,” JAMA Health Forum, vol. 2, no. 9, p. e212861, Sep. 2021, doi: 10.1001/jamahealthforum.2021.2861.

[23] R. M. Myerson, R. D. Tucker-Seeley, D. P. Goldman, and D. N. Lakdawalla, “Does Medicare Coverage Improve Cancer Detection and Mortality Outcomes?,” Journal of Policy Analysis and Management, vol. 39, no. 3, pp. 577–604, 2020, doi: 10.1002/pam.22199.

[24] J. Semprini, “Examining the ‘Medicare Effect’ on Distant-Stage Cancer Diagnoses by Site, Gender, and Rurality,” vol. 6, no. 1, Art. no. 1, Jul. 2023, doi: 10.29024/jsim.171.

[25] J. Sun, M. C. Perraillon, and R. Myerson, “The Impact of Medicare Health Insurance Coverage on Lung Cancer Screening.,” Med Care, vol. 60, no. 1, pp. 29–36, Jan. 2022, doi: 10.1097/MLR.0000000000001655.

[26] E. C. Paulson et al., “National Cancer Institute Designation Predicts Improved Outcomes in Colorectal Cancer Surgery,” Annals of Surgery, vol. 248, no. 4, p. 675, Oct. 2008, doi: 10.1097/SLA.0b013e318187a757.

[27] A. E. Anderson, K. A. Henry, N. J. Samadder, R. M. Merrill, and A. Y. Kinney, “Rural vs Urban Residence Affects Risk-Appropriate Colorectal Cancer Screening,” Clin Gastroenterol Hepatol, vol. 11, no. 5, pp. 526– 533, May 2013, doi: 10.1016/j.cgh.2012.11.025.

[28] C. L. Odahowski, W. E. Zahnd, and J. M. Eberth, “Challenges and Opportunities for Lung Cancer Screening in Rural America,” Journal of the American College of Radiology, vol. 16, no. 4, pp. 590–595, Apr. 2019, doi: 10.1016/j.jacr.2019.01.001.

[29] A. M. Cole, J. E. Jackson, and M. Doescher, “Colorectal Cancer Screening Disparities for Rural Minorities in the United States,” J Prim Care Community Health, vol. 4, no. 2, pp. 106–111, Apr. 2013, doi: 10.1177/2150131912463244.

[30] G. LeBlanc, I. Lee, H. Carretta, Y. Luo, D. Sinha, and G. Rust, “Rural-Urban Differences in Breast Cancer Stage at Diagnosis,” Women’s Health Reports, vol. 3, no. 1, pp. 207–214, Dec. 2022, doi: 10.1089/whr.2021.0082.

[31] J. Orwat, N. Caputo, W. Key, and J. De Sa, “Comparing Rural and Urban Cervical and Breast Cancer Screening Rates in a Privately Insured Population,” Soc Work Public Health, vol. 32, no. 5, pp. 311–323, 2017, doi: 10.1080/19371918.2017.1289872.

[32] M. Alyabsi, J. Meza, K. M. M. Islam, A. Soliman, and S. Watanabe-Galloway, “Colorectal Cancer Screening Uptake: Differences Between Rural and Urban Privately-Insured Population,” Frontiers in Public Health, vol. 8, 2020, Accessed: Sep. 14, 2023. [Online]. Available: https://www.frontiersin.org/articles/10.3389/fpubh.2020.532950

[33] J. T. McDaniel et al., “Rural–urban disparities in colorectal cancer screening among military service members and Veterans,” Journal of Military, Veteran and Family Health, vol. 5, no. 1, pp. 40–48, Apr. 2019, doi: 10.3138/jmvfh.2018-0013.

[34] N. Theodoropoulos, H. Xie, Q. Wang, C. Wen, and Y. Li, “Rural-urban differences in breast and colorectal cancer screening among US women, 2014-2019,” Rural Remote Health, vol. 22, no. 3, p. 7339, Sep. 2022, doi: 10.22605/RRH7339.

[35] A. M. Cole, J. E. Jackson, and M. Doescher, “Urban–rural disparities in colorectal cancer screening: cross-sectional analysis of 1998–2005 data from the Centers for Disease Control’s Behavioral Risk Factor Surveillance Study,” Cancer Med, vol. 1, no. 3, pp. 350–356, Dec. 2012, doi: 10.1002/cam4.40.

[36] S. C. Curtin and M. R. Spencer, “Trends in Death Rates in Urban and Rural Areas: United States, 1999-2019,” NCHS Data Brief, no. 417, pp. 1–8, Sep. 2021.

[37] J. Baker et al., “Rural–Urban Disparities in Health Access Factors Over Time: Implications for Cancer Prevention and Health Equity in the Midwest,” Health Equity, vol. 6, no. 1, pp. 382–389, May 2022, doi: 10.1089/heq.2021.0068.

[38] J. A. Thompson et al., “The need to study rural cancer outcome disparities at the local level: a retrospective cohort study in Kansas and Missouri,” BMC Public Health, vol. 21, no. 1, p. 2154, Nov. 2021, doi: 10.1186/s12889-021-12190-w.

[39] K. R. Yabroff, X. Han, J. Zhao, L. Nogueira, and A. Jemal, “Rural Cancer Disparities in the United States: A Multilevel Framework to Improve Access to Care and Patient Outcomes,” JCO Oncology Practice, vol. 16, no. 7, pp. 409–413, Jun. 2020, doi: 10.1200/OP.20.00352.

[40] T. C. Tucker et al., “Improving the Quality of Cancer Care in Community Hospitals,” Ann Surg Oncol, vol. 28, no. 2, pp. 632–638, Feb. 2021, doi: 10.1245/s10434-020-08867-y.

[41] J. A. Gale, “Twenty-five years of the Medicare Rural Hospital Flexibility Program: The past as prologue,” The Journal of Rural Health, 2023, doi: 10.1111/jrh.12754.

[42] M. Lahr, X. Santana, H. Parsons, N. Bean, and I. Moscovice, “Delivery of Cancer Screening and Treatment in Critical Access Hospitals | Flex Monitoring Team,” Flex Monitoring Team, Oct. 2022. Accessed: Sep. 14, 2023. [Online]. Available: https://www.flexmonitoring.org/publication/delivery-cancer-screening-and-treatment-critical-access-hospitals

[43] I. S. Moscovice et al., “Availability of cancer care services and the organization of care delivery at critical access hospitals,” Cancer Medicine, vol. 12, no. 16, pp. 17322–17330, 2023, doi: 10.1002/cam4.6337.

[44] J. A. Rizzo, “Has Medicare been a ‘bad deal’ for rural hospitals?,” J Rural Health, vol. 7, no. 5, pp. 599–617, 1991.

[45] L. D. Brown, “The Politics of Medicare and Health Reform, Then and Now,” Health Care Financ Rev, vol. 18, no. 2, pp. 163–168, 1996.

[46] N. McCall, J. Korb, A. Petersons, and S. Moore, “Reforming Medicare Payment: Early Effects of the 1997 Balanced Budget Act on Postacute Care,” Milbank Q, vol. 81, no. 2, pp. 277–303, Jun. 2003, doi: 10.1111/1468-0009.t01-1-00054.

[47] R. Slifkin, “Rural Hospital Closures, 1990-2000: Community Profiles and Economic Indicators Before and After the Event: Rural Health Research Project,” North Carolina Rural Health Research and Policy Analysis Center, Dec. 2004. Accessed: Sep. 15, 2023. [Online]. Available: https://www.ruralhealthresearch.org/projects/377

[48] Rural Health Information Hub, “Critical Access Hospitals (CAHs) Overview.” Accessed: Sep. 15, 2023. [Online]. Available: https://www.ruralhealthinfo.org/topics/critical-access-hospitals

[49] AHA, “Fast Facts on U.S. Hospitals, 2023.” Accessed: Sep. 15, 2023. [Online]. Available: https://www.aha.org/statistics/fast-facts-us-hospitals

[50] B. Wright, H.-Y. Jung, Z. Feng, and V. Mor, “Trends in Observation Care Among Medicare Fee-for-Service Beneficiaries at Critical Access Hospitals, 2007 – 2009,” J Rural Health, vol. 29, no. 0 1, pp. s1–s6, Aug. 2013, doi: 10.1111/jrh.12007.

[51] N. Natafgi, J. Baloh, P. Weigel, F. Ullrich, and M. M. Ward, “Surgical Patient Safety Outcomes in Critical Access Hospitals: How Do They Compare?,” J Rural Health, vol. 33, no. 2, pp. 117–126, Apr. 2017, doi: 10.1111/jrh.12176.

[52] A. A. Khaliq, E. Nsiah, N. H. Bilal, D. R. Hughes, and R. Duszak, “The Scope and Distribution of Imaging Services at Critical Access Hospitals,” Journal of the American College of Radiology, vol. 11, no. 9, pp. 857– 862, Sep. 2014, doi: 10.1016/j.jacr.2014.02.013.

[53] P. Hung et al., “Geographic disparities in residential proximity to colorectal and cervical cancer care providers,” Cancer, vol. 126, no. 5, pp. 1068–1076, Mar. 2020, doi: 10.1002/cncr.32594.

[54] C. F. Longacre, H. T. Neprash, N. D. Shippee, T. M. Tuttle, and B. A. Virnig, “Evaluating Travel Distance to Radiation Facilities Among Rural and Urban Breast Cancer Patients in the Medicare Population,” J Rural Health, vol. 36, no. 3, pp. 334–346, Jun. 2020, doi: 10.1111/jrh.12413.

[55] K. Weeks et al., “Rural disparities in surgical care from gynecologic oncologists among Midwestern ovarian cancer patients,” Gynecol Oncol, vol. 160, no. 2, pp. 477–484, Feb. 2021, doi: 10.1016/j.ygyno.2020.11.006.

[56] P. Hung et al., “Trends in Cancer Treatment Service Availability Across Critical Access Hospitals and Prospective Payment System Hospitals,” Medical Care, vol. 60, no. 3, pp. 196–205, Mar. 2022, doi: 10.1097/MLR.0000000000001635.

[57] S. Haneuse, F. Dominici, S.-L. Normand, and D. Schrag, “Assessment of Between-Hospital Variation in Readmission and Mortality After Cancer Surgical Procedures,” JAMA Netw Open, vol. 1, no. 6, p. e183038, Oct. 2018, doi: 10.1001/jamanetworkopen.2018.3038.

[58] K. Butts, “Geographic difference-in-discontinuities,” Applied Economics Letters, vol. 30, no. 5, pp. 615–619, Mar. 2023, doi: 10.1080/13504851.2021.2005236.

[59] J. Millán-Quijano, “Fuzzy di?erence in discontinuities,” Applied Economics Letters, vol. 27, no. 19, pp. 1552–1555, Nov. 2020, doi: 10.1080/13504851.2019.1696930.

[60] V. Grembi, T. Nannicini, and U. Troiano, “Do Fiscal Rules Matter?,” American Economic Journal: Applied Economics, vol. 8, no. 3, pp. 1–30, Jul. 2016, doi: 10.1257/app.20150076.

[61] NCI, SEER Incidence Data - SEER Data & Software. (Apr. 19, 2023). Accessed: Apr. 24, 2023. [Online]. Available: https://seer.cancer.gov/data/index.html

[62] Flex Monitoring Team, “Historical CAH Data.” Accessed: Nov. 06, 2022. [Online]. Available: https://www.flexmonitoring.org/historical-cah-data-0

[63] J. Ruhl, C. Callaghan, and N. Schussler, “Summary Staging Manual - 2018,” NCI, 2018. Accessed: Sep. 21, 2023. [Online]. Available: https://seer.cancer.gov/tools/ssm/index.html

[64] R. L. Siegel, K. D. Miller, N. S. Wagle, and A. Jemal, “Cancer statistics, 2023,” CA: A Cancer Journal for Clinicians, vol. 73, no. 1, pp. 17–48, 2023, doi: 10.3322/caac.21763.

[65] StataCorp, Stata Statistical Software. (2021). College Station, TX. StataCorp LLC. Accessed: May 04, 2023. [Online]. Available: https://www.stata.com/

[66] A. Abadie, S. Athey, G. W. Imbens, and J. M. Wooldridge, “When Should You Adjust Standard Errors for Clustering?*,” The Quarterly Journal of Economics, vol. 138, no. 1, pp. 1–35, Feb. 2023, doi: 10.1093/qje/qjac038.

[67] A. S. Venkataramani, J. Bor, and A. B. Jena, “Regression discontinuity designs in healthcare research,” BMJ, vol. 352, p. i1216, Mar. 2016, doi: 10.1136/bmj.i1216.

[68] R. L. Siegel, N. S. Wagle, A. Cercek, R. A. Smith, and A. Jemal, “Colorectal cancer statistics, 2023,” CA: A Cancer Journal for Clinicians, vol. 73, no. 3, pp. 233–254, 2023, doi: 10.3322/caac.21772.

[69] N. Howlader et al., “The Effect of Advances in Lung-Cancer Treatment on Population Mortality,” New England Journal of Medicine, vol. 383, no. 7, pp. 640–649, Aug. 2020, doi: 10.1056/NEJMoa1916623.

[70] T. Lu et al., “Trends in the incidence, treatment, and survival of patients with lung cancer in the last four decades,” Cancer Manag Res, vol. 11, pp. 943–953, Jan. 2019, doi: 10.2147/CMAR.S187317.

[71] K. C. Stoltzfus et al., “Impact of Facility Surgical Volume on Survival in Patients With Cancer,” Journal of the National Comprehensive Cancer Network, vol. 19, no. 5, pp. 495–503, Feb. 2021, doi: 10.6004/jnccn.2020.7644.

[72] N. E. Farrow et al., “Disparities in guideline-concordant treatment for node-positive, non–small cell lung cancer following surgery,” The Journal of Thoracic and Cardiovascular Surgery, vol. 160, no. 1, pp. 261-271.e1, Jul. 2020, doi: 10.1016/j.jtcvs.2019.10.102.

[73] J. M. McWilliams, “Don’t Look Up? Medicare Advantage’s Trajectory And The Future Of Medicare,” Health Affairs Forefront, Mar. 2022, doi: 10.1377/forefront.20220323.773602.

[74] N. C. Lee et al., “Implementation of the National Breast and Cervical Cancer Early Detection Program: the beginning,” Cancer, vol. 120 Suppl 16, no. 0 16, pp. 2540–2548, Aug. 2014, doi: 10.1002/cncr.28820.

[75] G. A. Brooks, J. R. Hoverman, and C. H. Colla, “The Affordable Care Act and Cancer Care Delivery,” Cancer J, vol. 23, no. 3, pp. 163–167, 2017, doi: 10.1097/PPO.0000000000000259.

[76] H. A. Moss, J. Wu, S. J. Kaplan, and S. Y. Zafar, “The Affordable Care Act’s Medicaid Expansion and Impact Along the Cancer-Care Continuum: A Systematic Review,” JNCI: Journal of the National Cancer Institute, vol. 112, no. 8, pp. 779–791, Aug. 2020, doi: 10.1093/jnci/djaa043.

[77] N. M. Mohr et al., “Outcomes Associated With Rural Emergency Department Provider-to-Provider Telehealth for Sepsis Care: A Multicenter Cohort Study,” Ann Emerg Med, vol. 81, no. 1, pp. 1–13, Jan. 2023, doi: 10.1016/j.annemergmed.2022.07.024.

[78] S. L. Schaefer, C. L. Mullens, and A. M. Ibrahim, “The Emergence of Rural Emergency Hospitals: Safely Implementing New Models of Care,” JAMA, vol. 329, no. 13, pp. 1059–1060, Apr. 2023, doi: 10.1001/jama.2023.1956.

