## Supplemental Files for "How did exposure to Critical Access Hospitals change Medicare’s effect on cancer survival?"

**Supplemental Exhibit 1: Cancer and Sex-Specific Summary Statistics**

| | | Distant Stage | | | Two-year Survival | | |
|---|---|---|---|---|---|---|---|
| | | **Aggregate** | **Female** | **Male** | **Aggregate** | **Female** | **Male** |
| All Sites | N | 1,042,339 | 497,860 | 544,479 | 1,245,952 | 568,231 | 677,721 |
| | mean | 23.5% | 24.3% | 24.0% | 72.2% | 69.7% | 68.3% |
| Lung | N | 150,792 | 67,129 | 83,663 | 164,487 | 73,341 | 91,146 |
| | mean | 55.8% | 54.5% | 57.5% | 30.2% | 31.4% | 24.4% |
| CRC | N | 111,761 | 53,014 | 58,747 | 115,634 | 54,821 | 60,813 |
| | mean | 21.6% | 19.3% | 19.9% | 76.7% | 73.2% | 73.0% |
| FB | N | - | 179,271 | - | - | 183,261 | - |
| | mean | - | 6.7% | - | - | 92.6% | - |
| Prostate | N | - | - | 194,238 | - | - | 247,417 |
| | mean | - | - | 3.3% | - | - | 95.5% |

N = analytic count of cancer cases. Mean = sample average for 65-year olds.

**Supplemental Exhibit 2: Visualizing the discontinuity at age 65 for all cancers combined**

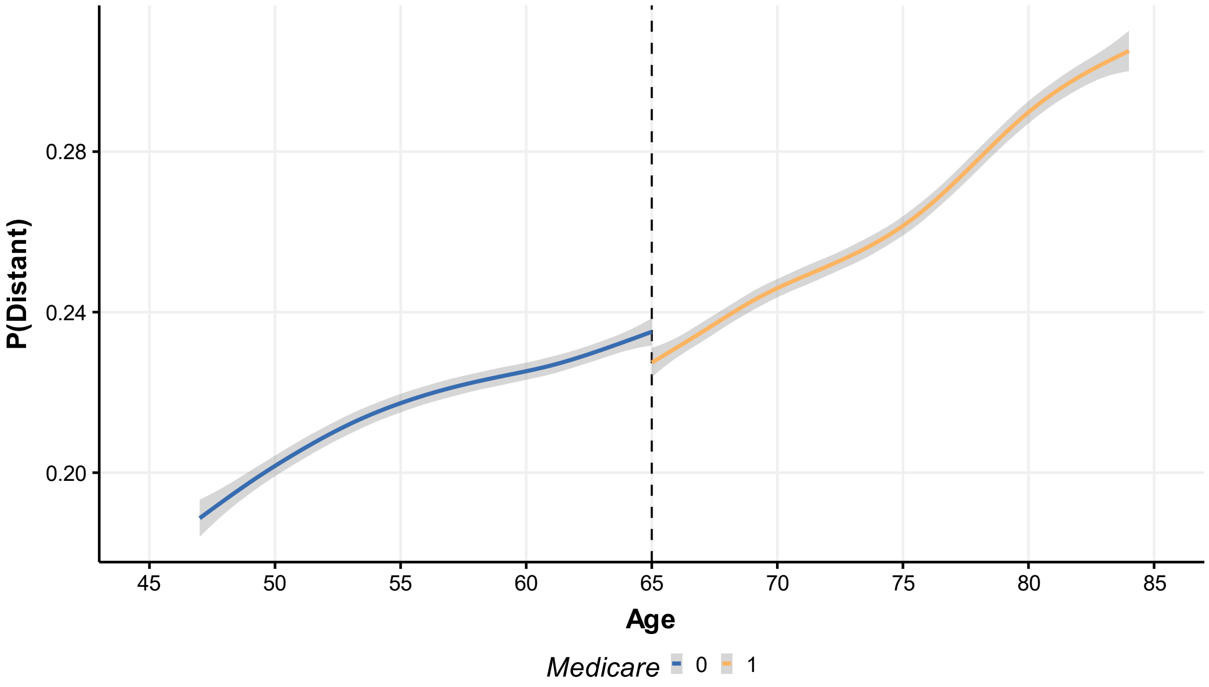


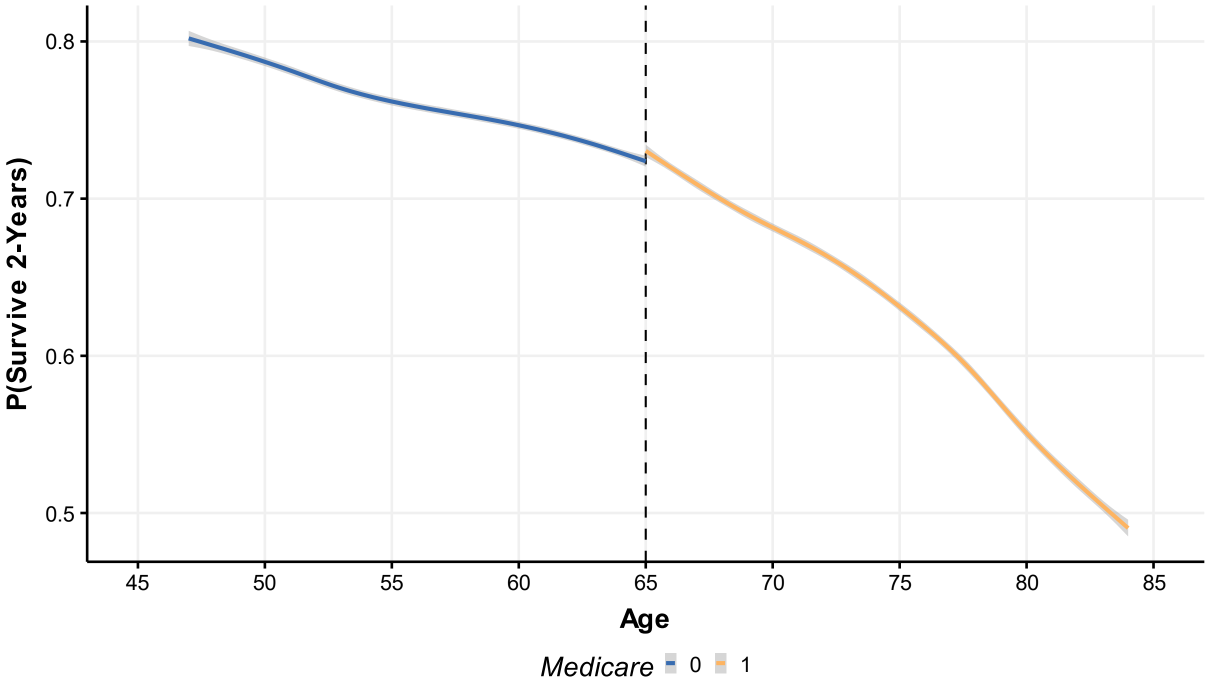


Supplemental Exhibit 2 plots the smoothed, local average outcome for all cancer cases (N=1,245,952). Lines represent the average probability at each age using locally estimated scatterplot smoothing method. Shaded gray area represents 95% Confidence Interval.

**Supplemental Exhibit 3: Differences-in-Discontinuities of a Distant Stage Diagnosis (All Cancers Combined)**

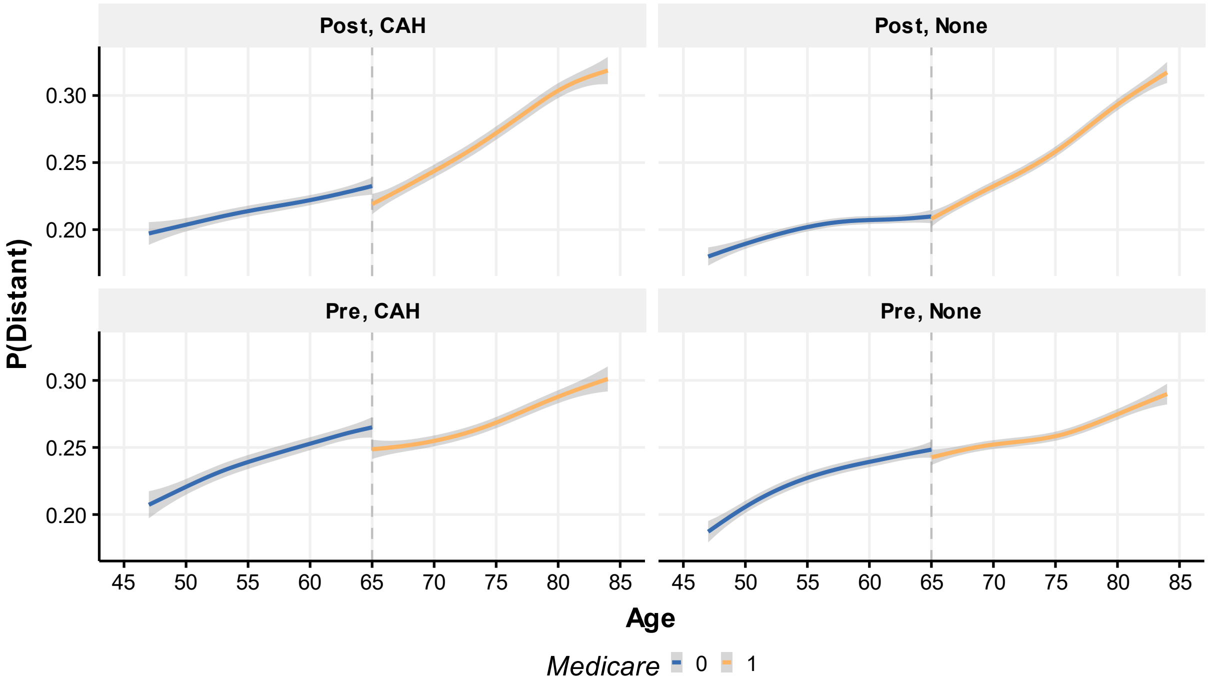


Supplemental Exhibit 3 plots the smoothed, local average probability of a distant stage diagnosis for all cancer cases (N=1,245,952). Lines represent the average probability at each age using locally estimated scatterplot smoothing method. Shaded gray area represents 95% Confidence Interval. Pre = 1993-1998, Post = 2006-2010. None = county did not have a CAH, CAH = county had at least one CAH.

**Supplemental Exhibit 4: Differences-in-Discontinuities of a Distant Stage Diagnosis (Lung, Colorectal, Female Breast, Prostate)**

**Lung Cancer**

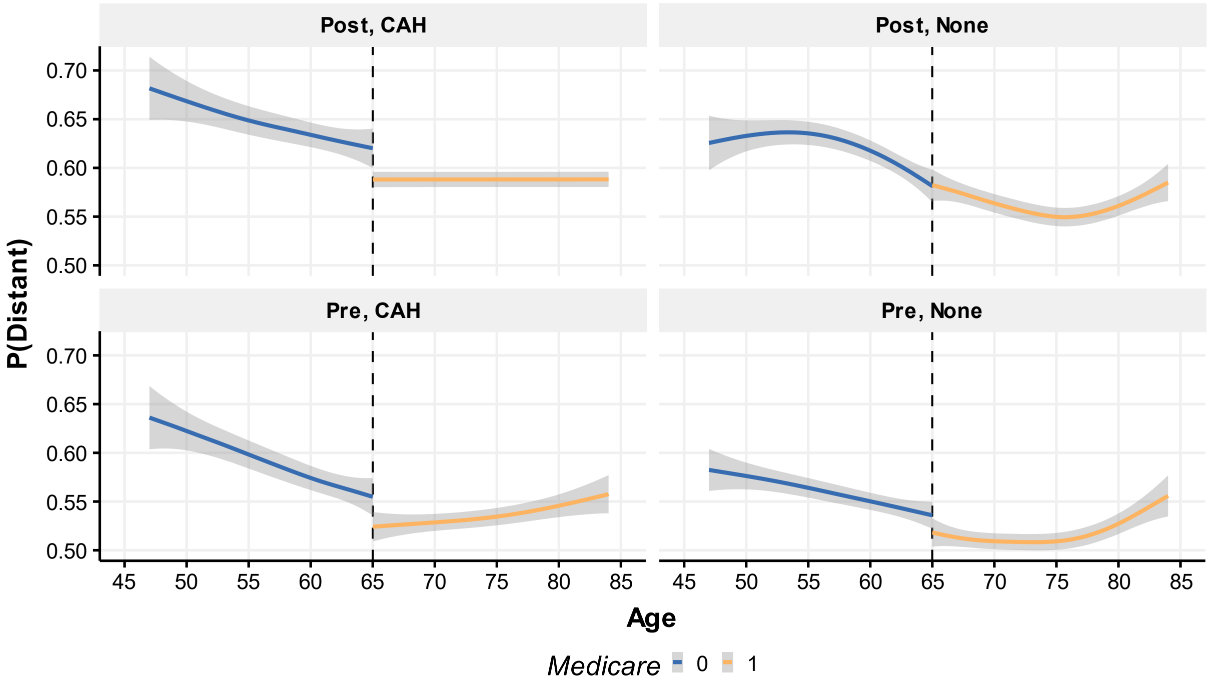


**Colorectal Cancer**

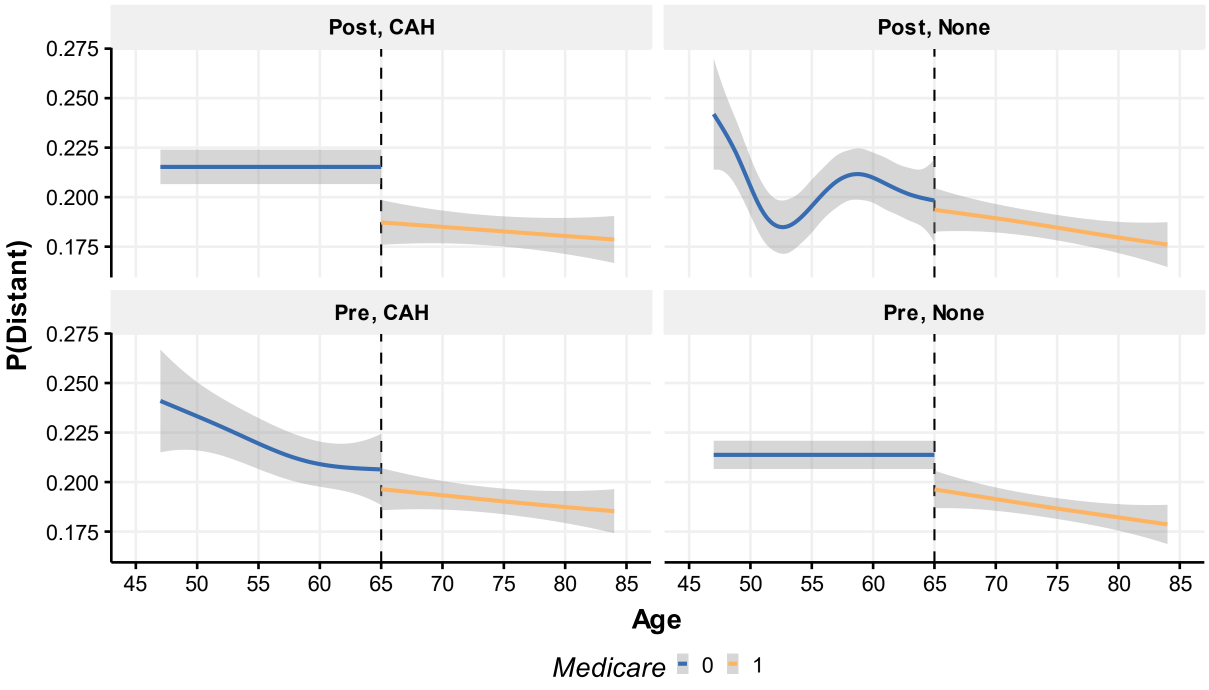

### Female Breast Cancer

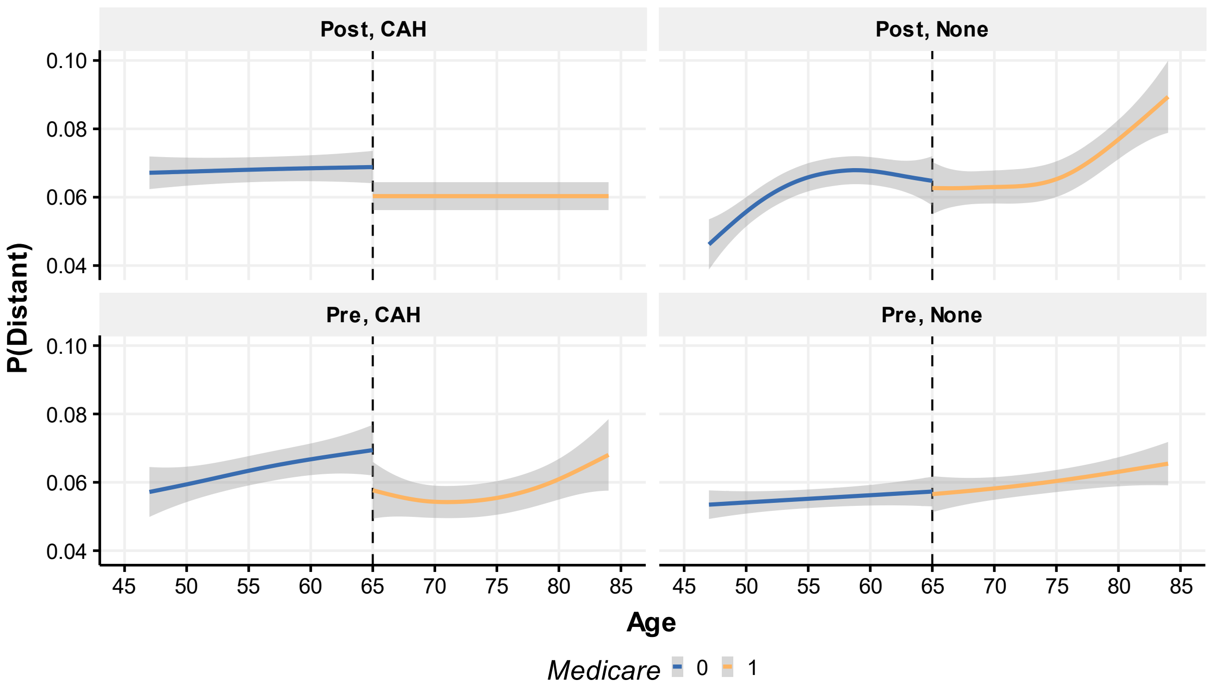


### Prostate Cancer

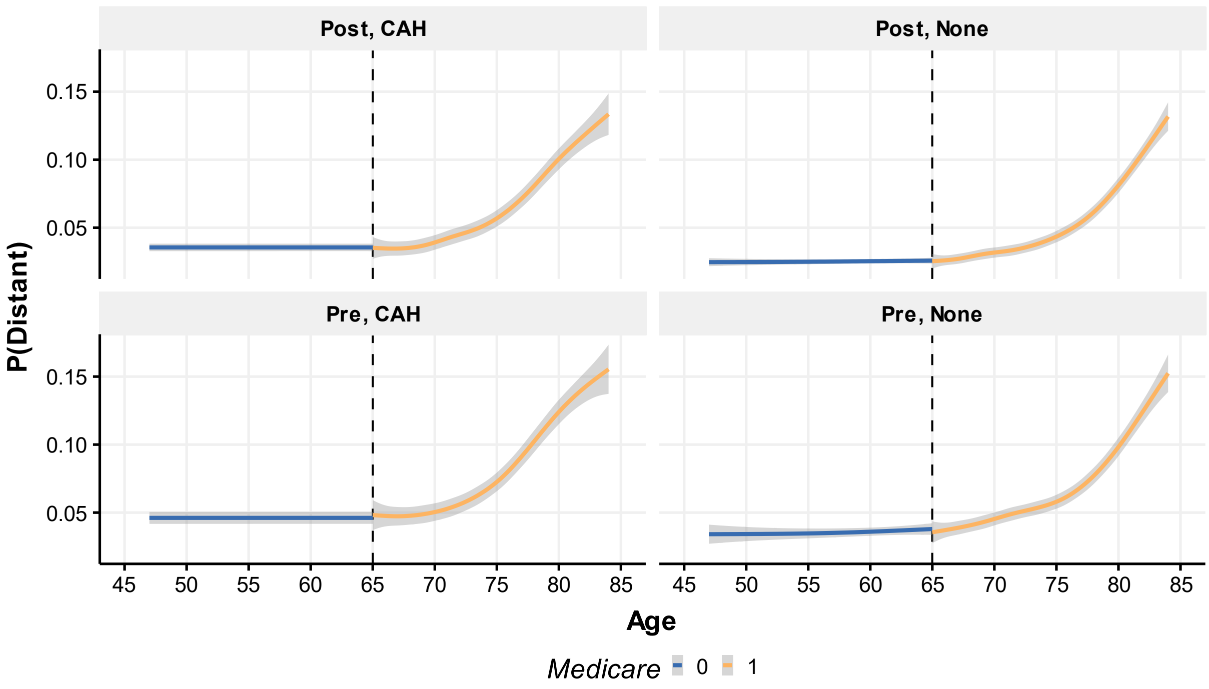


Supplemental Exhibit 3 plots the smoothed, local average probability of a distant stage diagnosis for lung, female breast, and prostate cancers. Lines represent the average probability at each age using locally estimated scatterplot smoothing method. Shaded gray area represents 95% Confidence Interval. Pre = 1993-1998, Post = 2006-2010. None = county did not have a CAH, CAH = county had at least one CAH.

**Supplemental Exhibit 5 – Alternative Specification Results – Distant Stage**

| | All Sites | Lung | Prostate | Female Breast | CRC |
|---|---|---|---|---|---|
| Primary | 0.004 | -0.003 | 0.001 | 0.014 | -0.035* |
| | (0.006) | (0.021) | (0.007) | (0.008) | (0.016) |
| Alt. 1 | -0.001 | -0.052 | -0.006 | -0.005 | -0.032 |
| | (0.010) | (0.033) | (0.009) | (0.012) | (0.023) |
| Alt 2. | 0.009 | 0.007 | 0.009 | 0.016 | -0.037* |
| | (0.006) | (0.023) | (0.005) | (0.009) | (0.016) |
| Placebo 1 | -0.001 | 0.031 | -0.023** | 0.010 | 0.000 |
| | (0.009) | (0.026) | (0.009) | (0.009) | (0.033) |
| Placebo 2 | -0.009 | 0.004 | -0.010 | 0.007 | 0.017 |
| | (0.010) | (0.021) | (0.012) | (0.017) | (0.016) |

Supplemental Exhibit 5 reports the estimated coefficient and standard error of the primary model and alternative specifications. Alt 1 = includes a set of quadratic age slopes. Alt 2 = includes a state-year, time-variant fixed-effect. Placebo 1 = tests for a discontinuity at age 55 cutoff. Placebo 2 = tests for a discontinuity at age 75 cutoff. Robust standard errors clustered at the county-level are reported in parentheses. * $p < 0.05$, $p < 0.01$

**Supplemental Exhibit 6: Alternative Specification Results – Two-Year Survival**

| | All Sites | Lung | Prostate | Female Breast | CRC |
|---|---|---|---|---|---|
| Primary | 0.002 | 0.019 | 0.019*** | -0.017* | 0.050** |
| | (0.005) | (0.015) | (0.005) | (0.008) | (0.016) |
| Alt. 1 | 0.012 | 0.058* | 0.019** | -0.015 | 0.079*** |
| | (0.008) | (0.023) | (0.007) | (0.011) | (0.021) |
| Alt. 2 | -0.005 | 0.013 | 0.016** | -0.019* | 0.046* |
| | (0.007) | (0.019) | (0.005) | (0.009) | (0.019) |
| Placebo 1 | -0.007 | -0.017 | 0.016 | -0.005 | -0.030 |
| | (0.010) | (0.029) | (0.008) | (0.008) | (0.031) |
| Placebo 2 | 0.004 | 0.003 | 0.013 | -0.015 | 0.041 |
| | (0.007) | (0.023) | (0.009) | (0.023) | (0.024) |

Supplemental Exhibit 6 reports the estimated coefficient and standard error of the primary model and alternative specifications. Alt 1 = includes a set of quadratic age slopes. Alt 2 = includes a state-year, time-variant fixed-effect. Placebo 1 = tests for a discontinuity at age 55 cutoff. Placebo 2 = tests for a discontinuity at age 75 cutoff. Robust standard errors clustered at the county-level are reported in parentheses. * $p < 0.05$ **, $p < 0.01$, *** $p < 0.001$
